# Associations of behavioral problems and white matter properties of the cerebellar peduncles in boys and girls born full term and preterm

**DOI:** 10.1101/2021.08.05.21261621

**Authors:** Machiko Hosoki, Lisa Bruckert, Lauren R. Borchers, Virginia A. Marchman, Katherine E. Travis, Heidi M. Feldman

**Author notes:** Corresponding author and contact information: Heidi M Feldman MD PhD, 3145 Porter Drive Mail Code 5395, Palo Alto, CA 94304, USA.

## Abstract

Accumulating evidence suggests that the role of cerebellum includes regulation of behaviors; Cerebellar impairment may lead to behavioral problems. Behavioral problems differ by sex: internalizing problems are more common in girls, externalizing problems in boys. Behavioral problems are also elevated in children born preterm (PT) compared to children born full-term (FT). The current study examined internalizing and externalizing problems in 8-year-old children in relation to sex, birth-group, FA of the cerebellar peduncles, and interactions among these predictor variables. Participants (*N*=78) were 44 boys (28 PT) and 34 girls (15 PT). We assessed behavioral problems via standardized parent-reports and FA of the cerebellar peduncles using deterministic tractography. Internalizing problems were higher in children born PT compared to children born FT (*p*=.032); the interaction of sex and birth-group was significant (*p*=.044). When considering the contribution of the mean-tract FA of cerebellar peduncles to behavioral problems, there was a significant interaction of sex and mean-tract FA of the ICP with internalizing problems; the slope was negative in girls (*p*=.020) but not boys. In boys, internalizing problems were only associated with mean-tract FA ICP in those born preterm (*p*=.010). We found no other significant associations contributing to internalizing or externalizing problems. Thus, we found sexual dimorphism and birth-group differences in the association of white matter metrics of the ICP and internalizing problems in school-aged children. The findings inform theories of the origins of internalizing behavioral problems in middle childhood and may suggest approaches to treatment at school age.

---

Behavioral problems in childhood are an important public health concern because of their high prevalence and contribution to short- and long-term adverse outcomes [1]. Behavioral problems encompass internalizing and externalizing problems. Internalizing problems refer to anxiety, fear, shyness, somatic complaints, withdrawal, and depression [2,3]. Externalizing problems refer to noncompliance, aggression, and destructiveness, also known as conduct problems [4]. Both types of behavioral problems typically begin in childhood and may persist as children develop [2,4,5]. Recent estimates from the National Survey of Children’s Health report prevalence rates among children ages 3 to 17 years at 3.2% for depression, 7.1% for anxiety, and 7.4% for externalizing behavioral or conduct problems [6]. Behavioral problems in childhood often negatively impact school performance and social relationships and may lead to long-term negative outcomes [1].

The cerebellum has been implicated in the neurobiological basis of behavioral problems. The cerebellum is situated in the posterior fossa, below the cerebral hemispheres, and connects to the cerebrum and to the spinal cord via three major white matter fiber bundles: the superior (SCP), middle (MCP), and inferior cerebellar peduncles (ICP). In cerebellar gray matters, investigations using functional MRI and positron emission tomography (PET) have suggested that cerebellar dysfunction is associated with behavioral disorders, including major depressive disorder, mania, obsessive-compulsive disorder, and autism [7–10]. In cerebellar white matters, investigation using diffusion MRI in young adults by Romer et al showed significant negative associations between fractional anisotropy (FA) of the SCP, the white matter bundle connecting the cerebellum to the frontal lobes, and behavioral problems [7].

Behavioral problems are known to differ in boys and girls: internalizing problems are more prevalent in girls than boys and become increasingly common in adolescence [3,5], while externalizing problems are more prevalent in boys than girls and may be observed early in childhood [11,12]. In addition, girls are more likely to show consistently elevated trajectories while boys may show variable trajectories as they grow older [13].

Sexual dimorphism is anatomically observed in the cerebellum, [14]; total cerebellar volume is greater in males than females, with differences gradually increasing during adolescence [15]. Borchers et al. [16] evaluated sexual dimorphism in the context of cerebellar white matter and behavioral problems. They found that lower FA of the SCP predicted increasing internalizing and externalizing problems only in adolescent girls [16]. They reported a similar pattern between FA of the ICP and externalizing problems. These findings suggest that cerebellar white matter properties are associated with behavioral problems in adolescence and there may be sexual dimorphism at this age. A gap in the literature is whether similar associations would be found in school-aged children, closer to the initial onset of behavioral problems.

Children born preterm are at risk for behavioral problems. Social-emotional and behavioral problems may be evident even before children turn 2 years of age [17,18]. They continue to exhibit behavioral problems through school-age [19] and into adulthood [13]. Preterm birth can cause cerebellar impairment [20], which in turn can lead to long-term adverse outcomes, such as motor disorders and behavioral dysfunction [21,22]. For example, volume of lateral cerebellar lobes and vermis were reduced in adolescents born before 33 weeks of gestation compared to adolescents born at term [23]. In the same study, decreased lateral cerebellar volume was correlated with reduced cerebral white matter volume, and was associated with reduced executive, language, and visuo-spatial skills [23]. In another study of young children, history of prematurity with isolated cerebellar hemorrhage was associated with more autistic behaviors and internalizing problems compared to children born preterm without cerebellar injury [24]. These studies suggest that cerebellar characteristics may play a role in behavioral problems in children born preterm. A gap in the literature is whether behavioral problems in children born preterm would be associated with microstructural characteristics of the cerebellar peduncles, the white matter pathways connecting the cerebellum to the cerebrum and spinal cord.

The current study aimed to address these gaps in the literature by investigating behavioral problems in relation to cerebellar white characteristics in school-aged children born at term or preterm. We divided behavioral problems into internalizing problems and externalizing problems because we anticipated that associations may be different in terms of neuropathological, and behavioral mechanisms. Our first aim was to evaluate whether behavioral problems would differ in school-aged girls and boys and in children born at term and preterm. We hypothesized that we would find higher levels of internalizing problems in girls as compared to boys and that we would observe higher levels of externalizing problems in boys. We also hypothesized that both internalizing and externalizing problems would be elevated in children born preterm compared to the children born at term. Our second aim was to evaluate whether mean-tract FA of the cerebellar peduncles would be associated with internalizing and externalizing problems, after considering sex and birth group. Given that cerebellum is associated with behavioral problem, we hypothesized that there will be associations between behavioral problems. Our third aim was to evaluate whether the association between behavioral problems and mean-tract FA of the cerebellar peduncles are moderated by sex or birth-group. Given sexual dimorphism in the nature and longitudinal emergence of behavioral problems, we hypothesized that sex [16] would moderate the associations between behavioral problems and mean-tract FA of the cerebellar peduncles. Given elevated rates of behavioral problems in children born preterm [18,25], we also hypothesized that birth-group would moderate the associations of mean-tract FA and behavioral problems.

## Materials and Methods

### Participants

The participants were boys and girls enrolled in a longitudinal study of reading skills and white matter characteristics in children born preterm and at term. The general characteristics of children enrolled have been described in previous studies [26–28]. Children were recruited from the San Francisco Bay Area from 2012 to 2015 and were followed for 2 years, from age 6 to 8 years. Preterm birth was defined as gestational age ≤ 32 weeks at birth and full term birth was defined as gestational age ≥ 37 weeks or birth weight ≥ 2500 grams. We defined children born ≤ 32 weeks as children born preterm, because very preterm birth increases the risk for white matter injury [29] and behavioral problems[25]. Exclusion criteria included hearing loss, visual impairment, history of neurological disorders, non-English speakers, and genetic disorders.

For this analysis, we included 8-year-old children whose parents completed the Child Behavior Checklist (CBCL), who underwent diffusion MRI, and whose scans were found to be free of motion artifacts and other technical problems. We decided to analyze 8 year old children, because we would capture more behavioral problems compared to 6 year old children given the nature and longitudinal emergence of behavioral problems [13]. The final sample included 78 children, 44 boys (26 children born at term) and 34 girls (15 children born preterm).

The experimental protocol was approved by the Stanford University Institutional Review Board. Written consent was obtained from a parent or a legal guardian and written assent was obtained from the participant. Children were compensated for participation. We collected demographic characteristics, including socioeconomic status using a modified Hollingshead Index [30], child sex, and birth-group.

### Assessments conducted at age 8

Parents completed the parent-reported CBCL/6-18, a questionnaire that indexes children’s behavioral problems [31]. The instrument generates broad band and narrow band scores. We focused on the main broad bands—internalizing and externalizing subscales—because broad bands have greater validity than narrow bands. Internalizing problems includes narrow band scores, which are anxious/depressed behavior, withdrawn-depressed behavior, and somatic complaints. Externalizing problems include narrow band scores, which are aggressive behavior and rule breaking behavior [31]. We analyzed raw scores of internalizing and externalizing problems because T scores account for age and sex.

### Diffusion MRI acquisition and analysis

MRI data were obtained on a 3T Discovery MR750 scanner (General Electric Healthcare, Milwaukee, WI, USA) equipped with a 32-channel head coil (Nova Medical, Wilmington, MsA, USA) at the Center for Cognitive and Neurobiological Imaging at Stanford University (https://cni.stanford.edu/). All children wore MRI-compatible headphones and watched a TV show of their choice during MRI scanning. We used a 3D fast-spoiled gradient (FSPGR) sequence (TR = 7.24 ms; TE = 2.78 ms; FOV = 230 mm × 230 mm; acquisition matrix = 256 × 256; 0.9 mm isotropic voxels; orientation = sagittal) to collect high resolution T1-weighted images. We used a dual-spin echo, echo-planar imaging sequence with full brain coverage (TR = 8300 ms; TE = 83.1 ms; FOV = 220 mm × 220 mm; acquisition matrix = 256 × 256; voxel size: 0.8594 mm× 0.8594 mm× 2 mm; orientation = axial) to collect diffusion MRI data with a b-value of 1000 s/mm^2^, sampling along 30 isotropically distributed diffusion directions. Three additional volumes were acquired at b = 0 at the beginning of each scan.

A detailed description of diffusion MRI preprocessing and tractography of the cerebellar peduncles is provided by Borchers et al. [32] and Bruckert et al. [33] based on the Automated Fiber-Tract Quantification methods described by Yeatman et al. [34]. The methods are summarized below.

We used the open-source software MrDiffusion (https://github.com/vistalab/vistasoft/tree/master/mrDiffusion) implemented in MATLAB 2014a (Mathworks, Natick, MA, United States) to preprocess the diffusion MRI data. In each child, we quantified head motion by measuring the amount of motion correction (in voxels) in all three planes of a diffusion volume relative to the previous volume. We then counted the number of volumes that had 1 or more voxels (voxel size = 2 mm^3^) of relative motion across the entire sample. Children who deviated from this mean by more than three standard deviations were excluded from analyses. (1 boy born at term was excluded.)

Using a rigid body alignment algorithm [35], we registered the b0 image to the participant’s anatomical T1-weighted image, which had been centered on the anterior commissure (AC) and aligned to the AC-PC plane. The combined transform that resulted from motion correction and alignment to the T1-weighted image was applied to the raw diffusion data (as well as the diffusion gradients [36]) and the transformed images were resampled to 2 × 2 × 2 mm^3^ isotropic voxels. We generated maps of fractional anisotropy (FA) using robust tensor fitting with outlier rejection [37].

We used the open-source toolbox Automated Fiber Quantification (AFQ)[34]; implemented in MATLAB 2014a to perform diffusion MRI tractography in a semi-automated fashion for each subject. First, whole-brain tractography was performed using a deterministic streamline tracking algorithm (STT)[38,39]. All voxels within a white matter mask with FA > 0.20 were defined as seed voxels. Tracking proceeded in all directions and was terminated in voxels with FA < 0.15 or when the angle of the principal diffusion direction of the current and next voxel was greater than 30°. Second, cerebellar white matter tracts were automatically segmented based on MNI template regions of interest (ROIs), as described by Travis et al.[28] and Bruckert et al. [33], warped into subject space. Third, cerebellar tracts were cleaned by removing streamlines that were either 4 standard deviations away from the core fiber of the tract, or that were more than 1 standard deviation longer or shorter than the mean length of the tract. Remaining streamlines were clipped to the same ROIs from which they were created, and we quantified FA at 30 equidistant locations, or ‘nodes’, along the tracts. The mean tract-FA was calculated by taking the average of all 30 nodes.

We visually inspected renderings of all segmented cerebellar tracts to ensure that the tracts conformed to known anatomical configurations. We successfully tracked the bilateral SCPs, MCP, and bilateral ICPs in most children. However, we could not segment the MCP, left ICP, or right ICP in 1, 7, and 8 children, respectively. We averaged the two hemispheres if there was no statistically significant lateralization between the left and right SCP and ICP based on Pearson correlation. For children who only had unilateral ICP tracked, we used the unilateral ICP mean-tract FA.

### Statistical analysis

All statistical analyses were conducted using SAS with statistical significance set at *p*<0.05. We conducted chi-squared tests to compare the proportion of children born at term and children born preterm within boys and girls. We compared age, SES, and mean-tract FA of cerebellar peduncles in the groups using independent samples t-test. We conducted a two-way ANOVA to evaluate if behavioral problems varied by sex or birth-group, and whether the interaction between sex and birth-group was statistically significant. If there was significant interaction, we ran post-hoc Tukey test in one-way ANOVA among four groups (boys born at term, boys born preterm, girls born at term, and girls born preterm) to determine which subgroup differed. We used Pearson correlation to describe strength of the association between internalizing and externalizing behavioral problems. We performed this analysis to determine whether to combine or separate internalizing and externalizing problems in subsequent analyses.

We conducted a series of hierarchical linear regression models to assess the contribution of mean-tract FA of each of the cerebellar peduncles to raw scores on the CBCL internalizing and externalizing scales at age 8. We standardized continuous variables; sex and birth group were dummy coded. First, we assessed the contribution of sex and birth-group to internalizing and externalizing scores. We then incorporated the contribution of the FA of the cerebellar peduncles to internalizing and externalizing scores. We evaluated the interaction term to determine if sex or birth-group moderated the association between behavioral problems and mean-tract FA of the cerebellar peduncles. Finally, if there was a significant interaction between behavioral problems and mean-tract FA of a particular cerebellar peduncle, we stratified the sample by sex to explore whether the association between the behavioral problems and the respective cerebellar peduncle was also moderated by birth-group. We used false discovery rate to adjust p-values within each cerebellar peduncle because the peduncles are anatomically independent from each other.

## Results

### Characteristics of participants

Table 1 presents the characteristics of the participants. There was no difference in proportions or the means in age, SES, behavioral problems, and mean-tract FA of cerebellar peduncles by birth group or sex. The correlation between internalizing and externalizing problems was significant in the entire sample (Supplement 1). However, the strength of these associations was weak to moderate, justifying separate analyses of internalizing and externalizing behavioral problems.

**Table 1:** Characteristics of the participants in terms of socioeconomic status, age, behavior problems, and tract FA by sex and birth-group

| Variable | Male<br>*M (SD) or N*<br>*N=44* |  | Female<br>*M (SD) or N*<br>*N=34* |  | Birth group<br>differences* | Sex<br>Differences* |
| --- | --- | --- | --- | --- | --- | --- |
|  | Term | Preterm | Term | Preterm | *p-value* | *p-value* |
| *N* | 16 | 28 | 19 | 15 | .086 |  |
| **Age (months)** | 74.9 (1.9) | 74.0 (2.0) | 74.1 (2.3) | 74.6 (2.7) | .624 | .953 |
| **SES** | 60.1 (7.8) | 52.5 (15.2) | 58.2 (11.4) | 57.2 (6.4) | .065 | .336 |
| **Mean-tract FA** |  |  |  |  |  |  |
| **SCP** | 0.44 (0.06) | 0.43 (0.04) | 0.45 (0.06) | 0.42 (0.05) | .199 | .553 |
| **MCP** | 0.55 (0.04) | 0.55 (0.06) | 0.53 (0.05) | 0.54 (0.02) | .808 | .133 |
| **ICP** | 0.45(0.04) | 0.45(0.03) | 0.44(0.03) | 0.45 (0.04) | .451 | .380 |

Note: SES: Socioeconomic status, SCP: Superior cerebellar peduncles, MCP: Middle cerebellar peduncle, ICP: Inferior cerebellar peduncles. \*Group differences were analyzed via independent.t-test.

Table 2 shows the results of the two-way ANOVA, which evaluated whether internalizing problems or externalizing problems differed by sex, birth-group, or the sex by birth-group interaction. Internalizing problems differed as a function of birth-group (*p*=.032). In addition, there was a significant interaction between sex and birth-group for internalizing problems (*p*=.044). Externalizing problems did not differ as a function of sex, birth-group, or the interaction of sex and birth-group.

**Table 2:** 2-way ANOVA result on internalizing problems raw score by sex and birth-group

| **Dependent** | **Factors** | **Sum of Squares** | **df** | **F-value** | ***p-value*** |
| --- | --- | --- | --- | --- | --- |
| **Internalizing problems** | Sex | 11.10 | 1 | 0.50 | .483 |
|  | Birth-group | 106.50 | 1 | 4.77 | **.032\*** |
|  | Sex x birth-group | 94.23 | 1 | 4.22 | **.044\*** |
| **Externalizing problems** | Sex | 60.55 | 1 | 2.28 | .135 |
|  | Birth-group | 8.96 | 1 | 0.34 | .563 |
|  | Sex x birth-group | 17.36 | 1 | 0.66 | .421 |

\* p<0.05 \*\* p<0.01.

### Characteristics of the cerebellar peduncles

We assessed mean-tract FA of the left and right SCP and left and right ICP. We found strong correlations between hemisphere (Supplementary table 2). Hence, we averaged the left and right SCP and ICP in all subsequent analyses.

### Associations of internalizing problems and mean-tract FA of the cerebellar peduncles

Table 3 shows the results of multiple regression models, which evaluated the association of mean-tract FA of the cerebellar peduncles with internalizing problems. Model 1A shows that sex and birth-group were not significantly associated with internalizing problems. Models 1B, 1C, and 1D demonstrated that there were no main effects of mean-tract FA of any of the cerebellar peduncles in association with internalizing problems.

**Table 3:**
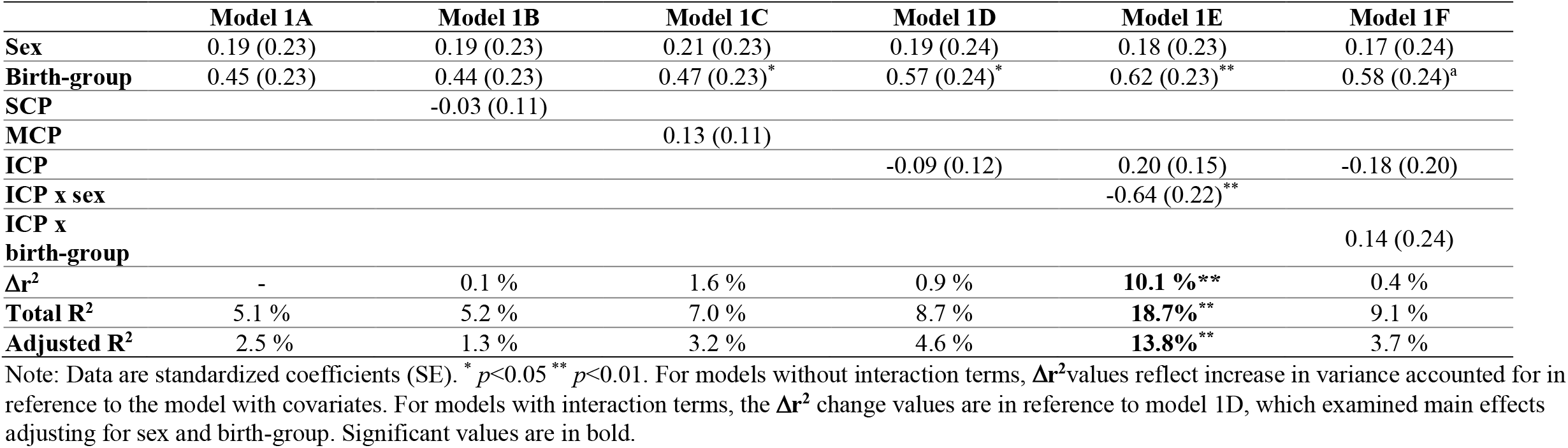
Contributions of mean-tract FA of ICP, sex, and birth-group for internalizing problems

Model 1E, which includes interaction term of sex and the mean-tract ICP FA, was statistically significant, accounting for 10.1% of the variance. This model survived FDR correction (adjusted *p* =.043). These results indicate that low ICP mean-tract FA was associated with higher internalizing problems in girls but not boys (Figure 1). This model remained significant, after excluding participants without bilateral ICP tracts (unadjusted interaction *p*=.011). The interaction of sex by tract did not contribute variance in internalizing problems for mean-tract FA of the SCP or MCP (Models 1R, 1T under supplementary Table 3). Models including the interaction of birth-group x tract were not statistically significant for the SCP, MCP, or ICP (for ICP: Model 1F under Table 3, SCP and ICP: Models 1S, 1U under supplementary table 3).

**Figure 1:**
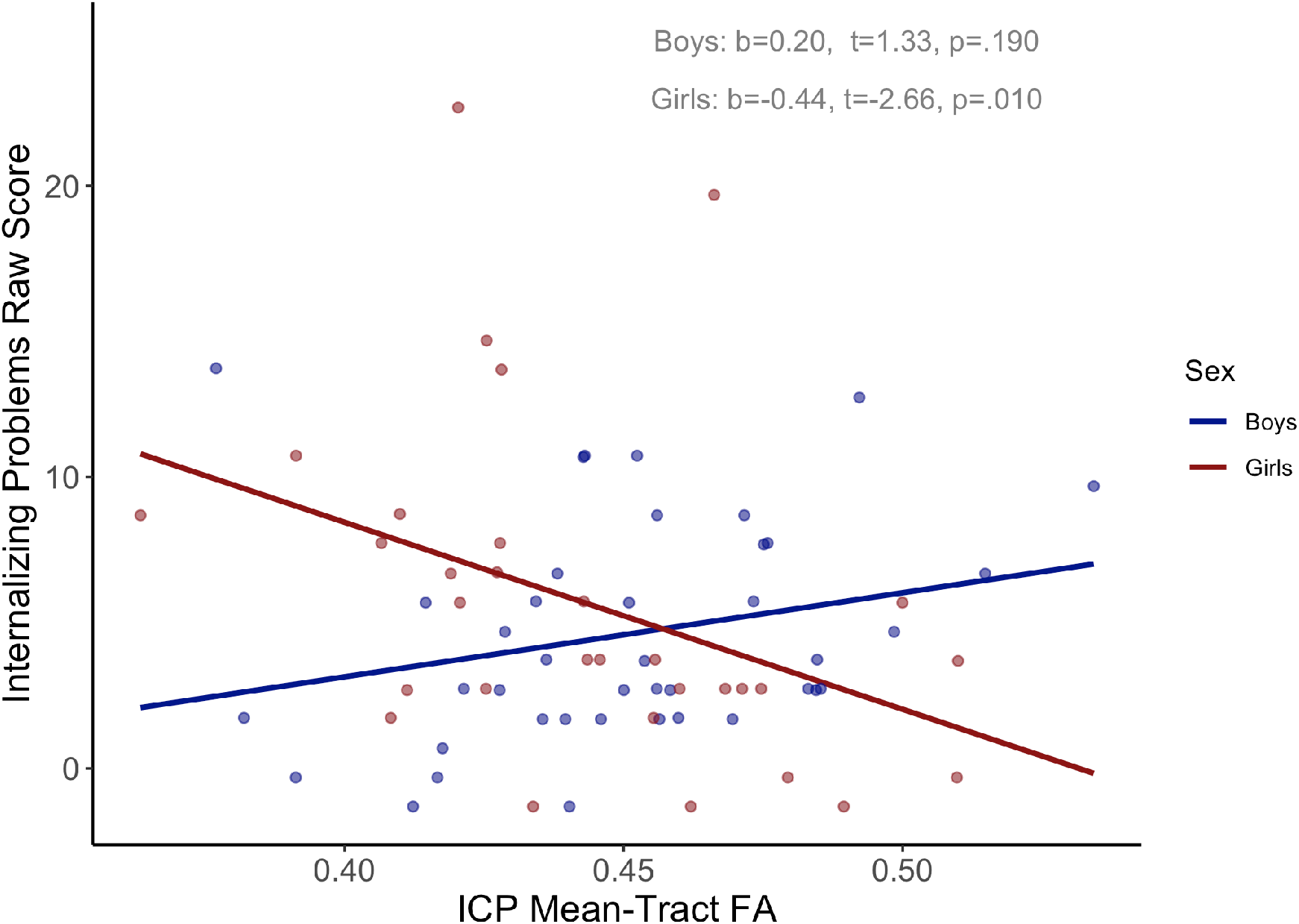
Lower mean-tract fractional anisotropy is associated with higher internalizing problems in girls. Note: Boy: blue circle, Girl: red circle. Covariate includes birth-group. ICP: Inferior cerebellar peduncle, FA: Fractional anisotropy

Because the association between internalizing problems and birth-group was moderated by sex, we further explored if birth-group moderated the association between internalizing problems and ICP mean-tract FA in boys and girls, respectively. In boys, association between internalizing problems and ICP mean-tract FA was moderated by birth-group, although the association was trending and did not fully meet statistical significance (interaction *t*=2.03, *p*=.050). Specifically, low ICP mean-tract FA was associated with low internalizing problems in boys born preterm but not boys born at term (Figure 2, boys). In girls, association between internalizing problems and ICP mean-tract FA did not differ by birth-group (interaction *t*=0.13, *p*=.901) (Figure 2, girls).

**Figure 2:**
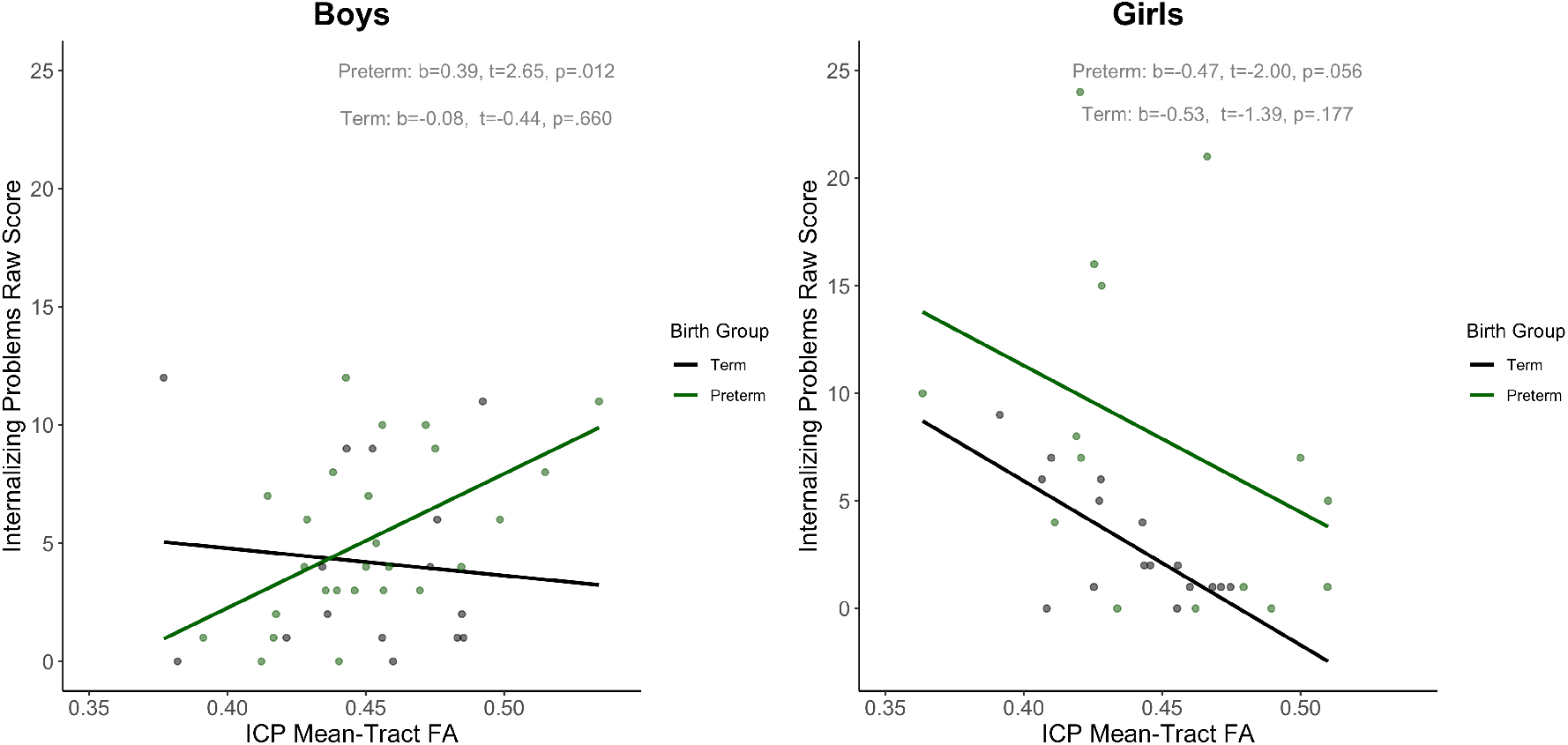
Associations between internalizing problems raw score and ICP mean-tract FA are moderated by birth-group in boys, but not girls.

### Associations of mean-tract FA of the cerebellar peduncles and externalizing problems

Supplementary table 4 demonstrates the results of a series of multiple regression models which explored the contribution of the mean-tract FA of the cerebellar peduncles to externalizing problems. Similar to the regressions relating to internalizing problems, sex and birth-group did not significantly contribute to externalizing problems. Models 2B, 2C, and 2D showed no association between mean-tract FA of the cerebellar peduncles and externalizing problems.

We analyzed the interaction with sex and mean tract FA of cerebellar peduncles in predicting externalizing problems. The models including the sex interaction were not statistically significant (models 2E, 2G, 2I for SCP, MCP, and ICP, respectively). We then considered the interaction with birth-group to the association between mean-tract FA of cerebellar peduncles and externalizing problems. The overall models including birth-group x tract were not statistically significant (models 2F, 2H, 2J for SCP, MCP, and ICP, respectively).

## Discussion

The purpose of the study was to investigate internalizing and externalizing behavioral problems in relation to sex, birth-group and mean-tract FA of the cerebellar peduncles in school-aged children. We were particularly interested in whether mean-tract FA of any of the cerebellar peduncles would contribute to individual differences in behavioral problems. Internalizing problems differed as a function of birth-group, and the association depended on sex. We found that internalizing problems were negatively associated with mean-tract FA of the ICP only in girls, not boys, and that internalizing problems were positively associated with mean-tract FA of the ICP only in boys born preterm, not boys born at term. This study, to our knowledge, is the first study that reported difference by sex and preterm birth in associations of behavioral problems and cerebellar peduncles in middle childhood.

The association of internalizing problems to sex and birth-group is consistent with other studies. Children born preterm are more prone to behavioral problems [40]. Girls have more internalizing problems than boys in general population[5,41], and girls born preterm have a higher susceptibility to internalizing problems than boys born preterm [42]. We did not find an association between externalizing problems, sex, and birth-group. While Indredavik et al [43] reported no sex differences in behavioral problems in adolescents born preterm, other studies report that boys born preterm have a higher susceptibility to externalizing problems than girls [42]. The lack of observed sex differences in externalizing problems may be due to sample size or other distinctive characteristics of this sample, such as high SES [44,45].

Our findings are consistent with previous studies that have implicated the cerebellar peduncles in behavioral problems. Romer et al. found that lower FA of the SCP was associated with more behavioral problems in young adults [7]. Borchers et al. highlighted sex-specific contributions of the cerebellar peduncles to behavioral problems, reporting that lower mean-tract FA of the SCP predicted increased internalizing and externalizing problems in adolescent girls [16] and that lower mean-tract FA of the ICP predicted increasing externalizing problems in adolescent girls. Our findings add that the association of behavioral problems and cerebellar peduncles is sexually dimorphic as early as school-age.

Interestingly, our finding of a negative association between internalizing problems and the mean-tract FA of ICP in girls differed from previous investigations that showed a negative association between internalizing problems and mean-tract FA of SCP [7,16]. These findings were interpreted based on the anatomy of the SCP. The SCP primarily contains efferent tracts and the peduncle is the main route for conveying cerebellar information to cerebral cortex [46]. The previous findings are consistent with studies that implicate the frontal lobe with behavior and emotion regulation [47].

The association of the ICP and internalizing problems may be related to anatomical and theoretical considerations. The ICP contains afferent and efferent fibers and is the main route for somatosensory information, such as proprioceptive stimuli, from muscle spindles and vestibular information via the brain stem to the brain [48]. Jang and Kwon [49] reported that ICP connects with the vestibular nucleus, reticular formation, pontine tegmentum with the posterior lobe of the cerebellum, which is hypothesized to play a role in emotional regulation [49]. Particularly, vermis of lobule VI is hypothesized as the “phylogenetically older cerebellar emotional processor” [40]. Hilber et al [50] have proposed that altered cerebellar and vestibular systems may change alter an individual’s sensation and perception; these alterations may result in false anticipation of motor commands and sensory feedback, which in turn compromise the individual’s ability to adjust to environmental stress and may give rise to a chronic anxiety state. Our findings appear to be consistent with proposed hypotheses; differences in the pattern of associations of mean-tract FA of the ICP may contribute to altering information to the posterior lobe of cerebellum and lead to a cascade of psychological changes associated with internalizing problems, such as anxiety.

In this study, we did not find associations between internalizing problems and mean-tract FA of the SCP, as previously reported [16]. One possible explanation is the difference in the age of participants. Cerebellar volume changes during middle childhood and adolescence, which suggests that there may be change of mean-tract FA[15]. As the cerebellum matures during this time period, there may be change in its functionality to behavioral problems within cerebellum. In children, internalizing problems may be related to connections of sensorimotor centers from body parts to the cerebellum. In adolescence and adults, internalizing problems may be related to connection between cerebellum and frontal areas of the cortex that are engaged in controlling emotions. Further research should verify the age differences in associations of internalizing problems and the cerebellar peduncles. Longitudinal data would be particularly useful in assessing whether the pattern of associations shifts within individuals. If these findings are verified, then there may be implications for therapeutic intervention; therapy for internalizing problems in young children may need to focus on assisting the child in detecting and interpreting somatosensory information whereas therapy for internalizing problems in adults or adolescents may need to focus on activating cognitive controls over emotional experiences, such as is emphasized in cognitive behavior therapy, an evidence-based psychological intervention for anxiety [51].

In boys, birth-group affected the association between internalizing problems and ICP mean-tract FA. While internalizing problems and ICP mean-tract FA was negatively associated in girls, the association was positive only in boys born preterm. Given that preterm birth often causes cerebellar damage [20], increased mean-tract FA could reflect compensation of myelinization from preterm birth [52], and increased mean-tract FA in children born may not reflect better white matter integrity [27]. However, we did not observe a similar association in girls born preterm. Future studies should verify our findings.

Though our study had many strengths, it also had three limitations. Our sample size was limited as we had 78 participants, reducing power to detect associations. We relied on parent-reported behavioral problems in the child, which may be less valid than self-reported behavioral problems, especially for internalizing problems, or objective measurements. Future analyses, which include larger sample sizes and use self-reported CBCL or observational measures are needed to confirm our findings. Finally, our participants were recruited on history of birth status, not clinical status of behavioral problems. Thus, future research is required to determine whether our findings generalize to cohorts of children with higher rates of elevated levels of behavioral problems than in the current sample.

## Conclusion

This study demonstrated negative associations between internalizing problems and cerebellar white matter properties in school-aged girls., such that lower mean-tract FA of ICP was associated with higher internalizing problems in girls. Our findings are in line with previous investigations that suggest that the cerebellar peduncle is related to emotional processing in adolescents [16] and adults [7], but further adds that child sex and preterm birth are important moderators of this relation. Externalizing problems were not associated with the cerebellar peduncles and there were no sex or birth-group interactions. Our findings suggest that internalizing and externalizing problems have different neurological bases. Future analysis may also consider analyzing change in the mean-tract FA of cerebellar peduncles over time and monitor response to therapy in children with behavioral problems.

## Data Availability

The data are de-identified, and stored in a secure and protected internal server, internally available at Stanford

## Acknowledgement

This work was supported by National Institutes of Health (NICHD grant RO1-HD069162 A and 2RO1-HD069150) and Health Resources and Services Administration Maternal Child Health Bureau (T77MC09796) to Heidi M Feldman. Machiko Hosoki is the Charles B. Woodruff Endowed Fellow Of the Maternal Child Health Research Institute, Stanford University.

## Tables

**Supplementary Table 4:** Contributions of mean-tract FA of ICP, sex, and birth-group for externalizing problems

|  | **Model 2A** | **Model 2B** | **Model 2C** | **Model 2D** | **Model 2E** | **Model 2F** |
| --- | --- | --- | --- | --- | --- | --- |
| **Sex** | 0.34 (0.23) | 0.34 (0.23) | 0.33 (0.24) | 0.32 (0.25) | 0.32 (0.25) | 0.29 (0.25) |
| **Birth-group** | 0.15 (0.23) | 0.15 (0.24) | 0.16 (0.24) | 0.20 (0.25) | 0.22 (0.25) | 0.21 (0.24) |
| **SCP** |  | -0.02 (0.12) |  |  |  |  |
| **MCP** |  |  | -0.03 (0.12) |  |  |  |
| **ICP** |  |  |  | -0.09(0.12) | 0.01 (0.17) | -0.35 (0.20) |
| **ICP x sex** |  |  |  |  | -0.22 (0.25) |  |
| **ICP x birth-group** |  |  |  |  |  | 0.40 (0.25) |
| **$\Delta r^2$** |  | 0.0 % | 0.0 % | 0.7 % | 1.1 % | 3.5 % |
| **Total R<sup>2</sup>** | 3.0 % | 3.0 % | 3.0 % | 3.8 % | 5.0 % | 7.4 % |
| **Adjusted R<sup>2</sup>** | 0.4 % | -0.9% | -1.0 % | -0.4 % | -0.7 % | 1.8 % |

Note: Data are standardized coefficients (SE). \* $p < 0.05$ \*\* $p < 0.01$ . For models without interaction terms, $\Delta r^2$ values reflect the increase in variance accounted for in reference to the model with covariates only. For models with interaction terms, the $\Delta r^2$ change values are in reference to model 2D, which examined main effects adjusting for sex and birth-group.

