## Supplementary tables for "Associations of behavioral problems and white matter properties of the cerebellar peduncles in boys and girls born full term and preterm"

Supplementary Table 1: Correlation between internalizing problems and externalizing problems

|  | **Pearson correlation coefficients** | ***p-value*** |
| --- | --- | --- |
| **Full Sample** | 0.42 | <.001 |
| **Boys** | 0.51 | <.001 |
| **Girls** | 0.36 | .034 |
| **Term** | 0.24 | .159 |
| **Preterm** | 0.56 | <.001 |

Supplementary Table 2: Measurement of mean-FA tract by left and right cerebellar superior/inferior peduncles

| **Variable** | | **Mean (SD)** | **Min** | **Max** | **Pearson correlation coefficients** | ***p-value*** |
| --- | --- | --- | --- | --- | --- | --- |
| **SCP** | Left | 0.44 (0.06) | 0.30 | 0.57 | 0.67 | <.001 |
|  | Right | 0.43 (0.05) | 0.30 | 0.58 |  |  |
| **ICP** | Left | 0.44 (0.03) | 0.36 | 0.53 | 0.72 | <.001 |
|  | Right | 0.46 (0.04) | 0.37 | 0.56 |  |  |

Note: SCP: Superior cerebellar peduncles, ICP: Inferior cerebellar peduncles.

Supplementary Table 3: Contributions of mean-tract FA of superior cerebellar peduncles, middle cerebellar peduncles, sex, and birth-group for internalizing problems

|  | **Model 1O** | **Model 1P** | **Model 1Q** | **Model 1R** | **Model 1S** | **Model 1T** | **Model 1U** |
| --- | --- | --- | --- | --- | --- | --- | --- |
| **Sex** | 0.19 (0.23) | 0.19 (0.23) | 0.21 (0.23) | 0.19 (0.23) | 0.18 (0.23) | 0.22 (0.24) | 0.20 (0.24) |
| **Birth-group** | 0.45 (0.23) | 0.44 (0.23) | 0.47 (0.23)^a^ | 0.44 (0.24) | 0.44 (0.23) | 0.46 (0.23) | 0.47 (0.23)^*^ |
| **SCP** |  | -0.03 (0.11) |  | -0.04 (0.16) | -0.01 (0.15) |  |  |
| **MCP** |  |  | 0.13 (0.11) |  |  | 0.09 (0.14) | 0.04 (0.18) |
| **SCP x sex** |  |  |  | 0.02 (0.23) |  |  |  |
| **SCP x birth-group** |  |  |  |  | -0.05 (0.24) |  |  |
| **MCP x sex** |  |  |  |  |  | 0.12 (0.26) |  |
| **MCP x birth-group** |  |  |  |  |  |  | 0.14 (0.23) |
| **Δr^2^** | - | 0.0% | 1.6 % | 0.0 % | 0.0 % | 0.3 % | 0.5 % |
| **Total R^2^** | 5.1 % | 5.2 % | 7.0 % | 5.2 % | 5.2 % | 7.3 % | 7.5 % |
| **Adjusted R^2^** | 2.5 % | 1.3 % | 3.2 % | 0.0 % | 0.0% | 2.2 % | 2.4 % |

Note: Data are standardized coefficients (SE). ^*^ p<0.05 ^**^ p<0.01. For models without interaction terms, **Δr^2^**values reflect increase in variance accounted for in reference to the model with covariates. For models with interaction terms, the **Δr^2^** change values are in reference to model 1P and IQ, which examined main effects adjusting for sex and birth-group. Significant values are in bold.

Supplementary Table 4: Contributions of mean-tract FA of cerebellar peduncles, sex, and birth-group for externalizing problems

|  | **Model 2A** | **Model 2B** | **Model 2C** | **Model 2D** | **Model 2E** | **Model 2F** | **Model 2G** | **Model 2H** | **Model 2I** | **Model 2J** |
| --- | --- | --- | --- | --- | --- | --- | --- | --- | --- | --- |
| **Sex** | 0.34 (0.21) | 0.34 (0.23) | 0.33 (0.24) | 0.32 (0.25) | 0.35 (0.23) | 0.32 (0.24) | 0.34 (0.24) | 0.34 (0.24) | 0.32 (0.25) | 0.29 (0.25) |
| **Birth-group** | 0.15 (0.23) | 0.15 (0.24) | 0.16 (0.24) | 0.20 (0.25) | 0.18 (0.23) | 0.14 (0.24) | 0.15 (0.24) | 0.16 (0.24) | 0.22 (0.25) | 0.21 (0.24) |
| **SCP** |  | -0.02(0.12) |  |  | -0.20(0.16) | 0.06 (0.15) |  |  |  |  |
| **MCP** |  |  | -0.03(0.12) |  |  |  | -0.05(0.14) | 0.05 (0.19) |  |  |
| **ICP** |  |  |  | -0.09(0.12) |  |  |  |  | 0.01 (0.17) | -0.35 (0.20) |
| **SCP x sex** |  |  |  |  | 0.36 (0.23) |  |  |  |  |  |
| **SCP x birth-group** |  |  |  |  |  | -0.20(0.24) |  |  |  |  |
| **MCP x sex** |  |  |  |  |  |  | 0.09 (0.26) |  |  |  |
| **MCP x birth-group** |  |  |  |  |  |  |  | -0.12(0.24) |  |  |
| **ICP x sex** |  |  |  |  |  |  |  |  | -0.22(0.25) |  |
| **ICP x birth-group** |  |  |  |  |  |  |  |  |  | 0.40 (0.25) |
| **Δr^2^** |  | 0.0 % | 0.0 % | 0.7 % | 3.1% | 0.9 % | 0.2 % | 0.3 % | 1.1 % | 3.5 % |
| **Total R^2^** | 3.0 % | 3.0 % | 3.0 % | 3.8 % | 6.2 % | 3.9 % | 3.2 % | 3.4 % | 5.0 % | 7.4 % |
| **Adjusted R^2^** | 0.4 % | -0.9% | -1.0 % | -0.4 % | 0.1 % | -1.4 % | -2.2 % | -2.0 % | -0.7 % | 1.8 % |

Note: Data are standardized coefficients (SE). ^*^ p<0.05 ^**^ p<0.01. For models without interaction terms, **Δr^2^**values reflect increase in variance accounted for in reference to the model with covariates. For models with interaction terms, the **Δr^2^** change values are in reference to model 2B, 2C, and 2D, which examined main effects adjusting for sex and birth-group. Significant values are in bold.
